# Identifying Clinical Diagnostic Trajectories Associated With Suicide Death Using Temporal Sequence Mining of Linked Claims and Mortality Data

**DOI:** 10.64898/2026.06.08.26355231

**Authors:** Anas Belouali, Christopher Kitchen, Emily Haroz, Harold Lehmann, Paul S. Nestadt, Holly C. Wilcox, Hadi Kharrazi

## Abstract

**Background:** Most approaches to suicide risk assessment consider clinical conditions as independent risk factors, potentially overlooking prognostic information in the order in which conditions accumulate. We applied temporal sequence mining to linked claims and mortality data to identify ordered clinical diagnostic trajectories associated with suicide death.

**Results:** The cohort included 3 647 059 insured Maryland residents aged 10 years or older with available claims records in the Maryland Suicide Data Warehouse from January 1, 2016, to December 31, 2020, among whom 768 suicide deaths were ascertained through medical examiner linkage. Sequential pattern mining of ICD-10-CM diagnoses grouped into Clinical Classifications Software Refined categories identified 89 221 candidate sequences, of which 1 816 remained significantly associated with suicide death in time-varying Cox models. Adjusted hazard ratios (AHRs) ranged from 2.4 to 134.1. Two-thirds of significant trajectories ended in physical conditions, and approximately half crossed from psychiatric to physical endpoints. Among suicide decedents, 62% were exposed to at least 1 significant sequence (median, 16 per case); median sequence duration was 18.7 months, and median time from completion to death was 13.1 months. In landmark analyses, among patients with depression who later developed suicidal ideation (n = 26 356), the path through anxiety, then anemia, was associated with higher risk (AHR, 4.6; 95% CI, 2.2-9.5), whereas the anxiety-only path was not (AHR, 1.3; 95% CI, 0.8-2.1). Among patients with anxiety who later developed hypertension (n = 149 215), the path through history of self-harm was associated with higher risk (AHR, 32.0; 95% CI, 16.6-61.6). Associations were generally consistent across sex and age.

**Conclusions:** Temporal ordering of clinical conditions may carry prognostic information for suicide death. Clinical trajectories incorporating physical illness within psychiatric sequences identified higher-risk groups. These findings suggest that opportunities for risk detection may extend beyond psychiatric settings and that suicide risk signals may be fragmented across care settings and not apparent within isolated encounters.

## Background

Suicide remains a leading cause of death in the United States, with more than 49 000 deaths annually [1]. Despite decades of research on identifying individual risk factors, clinical risk assessment for suicide remains imprecise [2–5]. Most approaches consider conditions as independent, static predictors and may miss prognostic information in how risk factors accumulate over time [6].

Evidence suggests that the temporal ordering of conditions may be meaningful for suicide risk. The relative timing of physical and psychiatric illness differentiated risk, with higher risk observed when the two occurred close together and when psychiatric disorder developed after physical illness [7]. However, whether longer diagnostic sequences of physical and psychiatric conditions carry differential risk for suicide has not been examined. Beyond physical-psychiatric timing, the order of psychiatric diagnoses themselves may also matter. For example, depression preceding a later diagnosis of bipolar disorder may represent early-stage bipolar illness, which is often more treatment-resistant than stable unipolar depression; antidepressant treatment in such patients may worsen mood instability and, in some cases, increase suicidal risk [8,9].

Data-driven approaches such as sequential pattern mining can identify recurrent, ordered diagnosis sequences in longitudinal data. Such methods capture patterns of condition accumulation that cross-sectional analyses can miss. They may also reveal temporal patterns that reflect evolving clinical presentations or care needs. Prior work has applied sequential pattern mining in electronic health records and claims data to study disease progression, adverse events, and treatment pathways [10–12]. In this study, we applied sequential pattern mining to statewide claims data to identify ordered sequences of clinical conditions associated with suicide death in a large insured population and to test whether, among individuals sharing the same starting and endpoint conditions, intermediate diagnostic paths were associated with differential subsequent suicide risk.

## Methods

This retrospective cohort study was approved by the Johns Hopkins Bloomberg School of Public Health Institutional Review Board with a waiver of informed consent. We followed the Strengthening the Reporting of Observational Studies in Epidemiology (STROBE) reporting guideline. The overall analytic workflow is shown in Fig. 1.

**Fig. 1.**
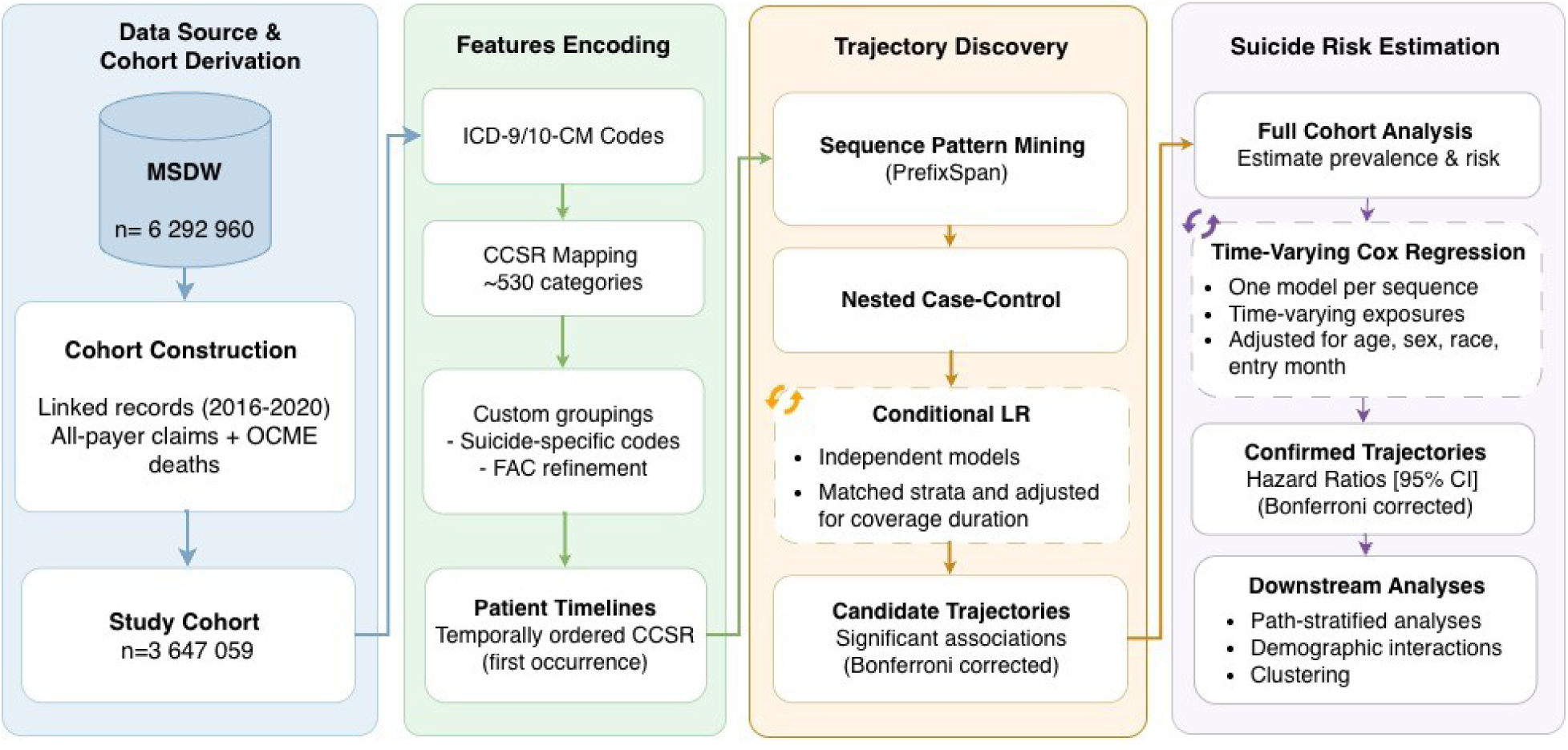
Analytic workflow for trajectory discovery and validation. Abbreviations: CCSR, Clinical Classifications Software Refined; CI, confidence interval; FAC, factors influencing health status; ICD-CM, International Classification of Diseases, Clinical Modification; LR, logistic regression; MSDW, Maryland Suicide Data Warehouse; OCME, Office of the Chief Medical Examiner.

### Data Source and Study Population

We used the Maryland Suicide Data Warehouse (MSDW), which links statewide health and mortality data for suicide research [13]. The analytic cohort was derived from all-payer claims provided by the Maryland Health Care Commission. Individuals were eligible if they were aged 10 years or older and had medical coverage between January 1, 2016, and December 31, 2020. Suicide death was ascertained through linkage with the Maryland Office of the Chief Medical Examiner records via the Chesapeake Regional Information System for Patients. Among 3 647 059 insured individuals, 768 died by suicide (Additional file 1: Fig. S1).

### Sequence Representation and Mining

A sequence was defined as an ordered list of clinical conditions based on the date of first recorded occurrence for each condition. Sequences reflect mined subsets of recorded conditions rather than complete patient histories, and the terms sequence and trajectory are used interchangeably. International Classification of Diseases, Tenth Revision, Clinical Modification (ICD-10-CM) diagnosis codes were mapped using the Clinical Classifications Software Refined (CCSR), version 2025.1, which aggregates ICD-10-CM codes into clinically meaningful categories [14]. We refer to these CCSR categories as clinical conditions. Custom groupings were created for suicide- and self-harm-related codes, and Z-codes were reclassified to exclude purely administrative encounters and separate nonspecific categories into more clinically specific subgroups. For readability, shortened clinical labels are used (eg, anxiety for anxiety and fear-related disorders). Reclassification details are provided in Additional file 1: Tables S1–S2.

For sequence mining, we retained only the first recorded occurrence of each condition per patient, yielding 71 million diagnosis records. We applied sequential pattern mining using the PrefixSpan algorithm [15], which expands sequences only when shorter patterns meet a minimum frequency threshold, avoiding the need to test every possible ordering. Sequences were retained only if observed in at least 11 suicide decedents, balancing statistical power and interpretability (Additional file 1: Fig. S2). The algorithm identifies both single-condition patterns (eg, depression, anxiety) and multi-condition trajectories (eg, depression → anxiety) that meet the frequency threshold among suicide decedents. The arrow (→) in sequences denotes temporal order based on the date of first recorded occurrence, not necessarily disease onset or causal relationship.

### Handling Same-Day Events

The presence of multiple diagnostic codes recorded on the same calendar date creates challenges for temporal pattern mining. During sequence mining, we imposed a deterministic intra-date ordering by sorting same-day codes alphabetically by CCSR category. This avoided the combinatorial explosion of enumerating all possible intra-date permutations, which would create multiple timelines per patient and make sequencing computationally infeasible. Although this may omit some valid but differently ordered same-day patterns, it ensures reproducibility and consistency when timestamps within a day are unavailable.

To mitigate potential loss of sensitivity introduced during the mining phase, we relaxed the ordering constraint during downstream patient-level sequence matching. Specifically, when identifying patients who completed a given mined sequence, we treated all CCSR codes recorded on the same date as an unordered set. A patient was considered to have completed a target sequence if all elements appeared in the correct sequential order across their longitudinal history, regardless of the order in which codes occurred on a shared date. This approach preserved temporal integrity across distinct dates while accounting for real-world coding practices.

### Statistical Analysis

We used a 2-stage analytic approach to identify sequences associated with suicide death and evaluate whether, among individuals sharing the same start and endpoint diagnoses, intermediate diagnostic paths were associated with differential suicide risk.

#### Sequence Discovery and Screening

Candidate sequences were screened in a matched discovery cohort. Of 768 suicide decedents, 694 had at least 1 recorded diagnosis and were included as cases in the matched discovery cohort. Each case was matched to 10 controls on sex, race and ethnicity, age (within 5 years), and calendar timing of insurance coverage (total N = 7 634). Matching used incidence density sampling to align observation windows and exposure opportunities [16]. All standardized mean differences were less than 0.10 (Additional file 1: Fig. S3 and Table S3). Associations were estimated using conditional logistic regression with Bonferroni correction for the number of candidate sequences tested (n = 89 221). Sequences meeting the corrected significance threshold were then evaluated in time-varying survival models in the full cohort.

#### Time-Varying Survival Models

Sequences identified in screening were evaluated in the full cohort using Cox proportional hazards models with time-varying exposures. We used a counting-process formulation with start and stop times to avoid immortal time bias [17,18]. Individuals entered the risk set at the start of observable medical coverage, and time was measured in days since cohort entry. Individuals contributed unexposed person-time until 30 days after sequence completion and exposed person-time thereafter. Suicide deaths during the lag were attributed to the unexposed group to reduce protopathic bias [19], in which diagnoses recorded shortly before death may reflect the terminal crisis. Age was modeled as time-varying and updated across intervals. Models were adjusted for sex, race and ethnicity, and stratified by entry year. Robust standard errors clustered at the individual level accounted for multiple intervals per person. Sequences that remained significant at a Bonferroni-corrected threshold (two-sided P < 0.05) in the time-varying models were considered confirmed.

#### Landmark Path Analyses

We conducted landmark analyses to test whether suicide risk differed across distinct diagnostic paths sharing the same origin and endpoint. We defined the origin as the first condition in a mined sequence, the endpoint as the last condition in the sequence, and the path as the intermediate conditions between them. The endpoint serves as the landmark and the index date for assessing subsequent suicide risk, but it does not represent the end of the patient’s full clinical timeline.

From the confirmed sequences, we selected those with the highest adjusted hazard ratios, clinical relevance as screening or intervention points, and enough outcome events across mutually exclusive paths to support stratified comparisons. For each origin-endpoint pair, we defined mutually exclusive paths and compared each specified path with the reference group comprising all other paths to the same endpoint.

Post-landmark suicide risk was estimated using Cox proportional hazards models. Follow-up began at the landmark diagnosis date plus the lag period and continued until suicide death, loss of coverage, or end of follow-up. Models were adjusted for age, sex, race and ethnicity, calendar year, observable duration, 12-month coverage completeness, and Charlson Comorbidity Index at trajectory start.

#### Sensitivity Analyses and Bias Mitigation

We conducted several sensitivity analyses to assess robustness to model specification and potential surveillance bias. These included alternative covariate adjustment strategies, additional control for sequence duration, and analyses stratified by sex and age.

We considered several potential sources of bias in the study design and analysis. Selection bias and unequal exposure opportunity were addressed through incidence density sampling in the discovery cohort. Immortal time bias was addressed by modeling sequences as time-varying exposures, and protopathic bias was reduced by applying a 30-day lag after sequence completion before exposure classification. Surveillance bias was evaluated through adjustment for observable follow-up duration and insurance coverage continuity, and through sensitivity analyses described above. However, residual confounding remains possible because claims data do not capture all relevant clinical factors.

Data analysis was performed from November 2024 to December 2025 using Python version 3.10 and R version 4.4.

## Results

Among 3 647 059 insured individuals with medical coverage from 2016 through 2020, 768 died by suicide (0.02%). Mean (SD) age at death was 50.9 (19.4) years (range, 10-99 years); 589 (76.7%) were male. Race was White for 462 decedents (60.2%), Black for 119 (15.5%), Asian for 31 (4.0%), and other or unknown for 156 (20.3%). Median follow-up was 4.3 years (IQR, 2.0-5.0) overall, 3.2 years (IQR, 2.0-4.3) among suicide decedents, and 4.3 years (IQR, 2.0-5.0) among controls.

### Sequence Discovery and Confirmation

Sequential pattern mining identified 89 221 candidate sequences meeting the minimum frequency threshold. In the matched discovery cohort, 1845 sequences were significantly associated with suicide death after Bonferroni correction. In time-varying Cox models of the full cohort, 1 816 sequences remained significantly associated with suicide death, of which 24 were single-condition associations (Additional file 1: Fig. S4). Depression was the most common starting condition, appearing in 52.7% of significant sequences, followed by suicidal ideation (21.6%) and anxiety (20.8%). Two-thirds of significant trajectories ended in physical conditions, and approximately half crossed from a psychiatric origin to a physical endpoint. Among suicide decedents, 62% were exposed to at least 1 significant sequence (median 16 per case; IQR, 3-81). Median sequence duration was 18.7 months (IQR, 8.2-28.4), and the median time from sequence completion to death was 13.1 months (IQR, 5.2-22.7). Across candidate sequences, longer trajectories were less common overall but more frequent among suicide decedents than controls (Additional file 1: Fig. S5). We organized confirmed sequences into 8 clinical domains: suicide attempt (SA), self-harm, suicidal ideation (SI), substance use, depression-to-metabolic dysfunction, psychosis-to-bipolar transition, depression-to-bipolar transition, and psychosocial adversity. Fig. 2 displays the 3 highest-risk sequences per domain alongside the most relevant single condition. Adjusted hazard ratios ranged from 2.4 to 134.1, with multistep sequences generally conferring greater risk than single diagnoses alone.

**Fig. 2.**
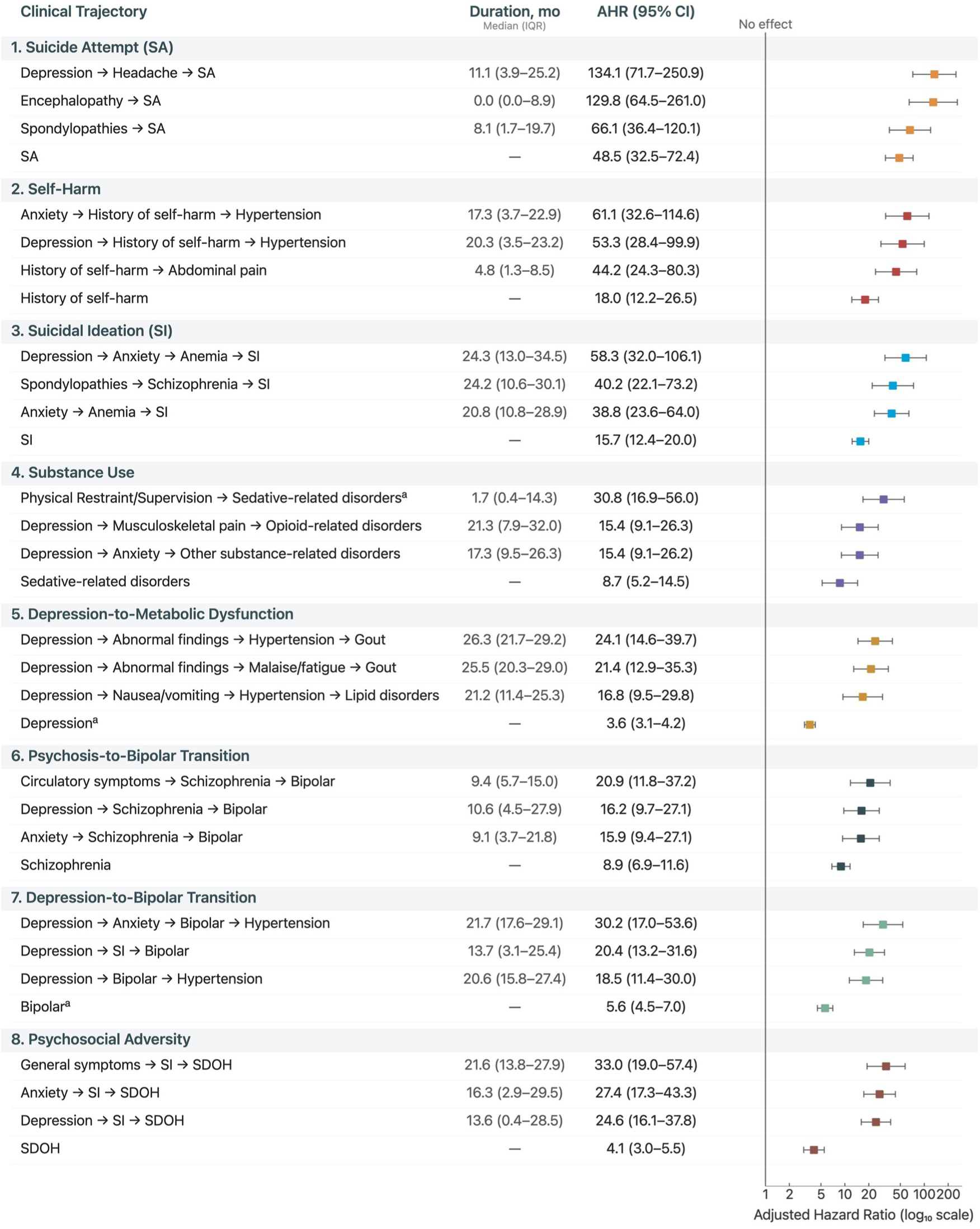
Selected sequences of clinical conditions associated with suicide death in the study population. Abbreviations: AHR, adjusted hazard ratio; CI, confidence interval; IQR, interquartile range; mo., months; SA, suicide attempt; SI, suicidal ideation; SDOH, social determinants of health. Arrows (→) indicate the order in which diagnoses were recorded. AHRs displayed on log₁₀ scale. Adjusted for age, sex, race/ethnicity, and cohort entry year. Representative trajectories from each domain are shown alongside single diagnoses as a reference. Duration is the median time in months from first to last diagnosis in the sequence, calculated among suicide decedents only. ^a^ Proportional hazards assumption violated; AHR represents time-averaged effect over follow-up.

### Landmark Path Analyses

Among individuals who shared the same origin and endpoint diagnosis, post-landmark suicide risk differed by intermediate diagnostic path (Table 1). Among patients with depression who later had an SA, paths through encephalopathy (AHR, 6.2; 95% CI, 2.3-16.8) and headache (AHR, 4.1; 95% CI, 1.7-10.0) were associated with higher risk than other paths to SA. Among patients with depression who later developed SI, the anxiety → anemia path was associated with higher risk (AHR, 4.6; 95% CI, 2.2-9.5), whereas the anxiety-only path was not (AHR, 1.3; 95% CI, 0.8-2.1). History of self-harm yielded the largest associations, with an AHR of 32.0 (95% CI, 16.6-61.6) among patients with anxiety who developed hypertension. Across cardiometabolic endpoints, physical-illness intermediates distinguished higher- from lower-risk paths; the nausea/vomiting → hypertension path to lipid disorders was elevated (AHR, 4.4; 95% CI, 2.3-8.4), whereas the hypertension-only path was not (AHR, 1.2; 95% CI, 0.8-1.8). For trajectories ending in social determinants of health (SDOH)-related codes, suicidal ideation as an intermediate was associated with elevated risk regardless of origin (AHRs, 6.4-14.2). Path-specific differences extended to bipolar transitions, psychosis-spectrum trajectories, and substance-related disorders. Cumulative incidence curves confirmed separation between high-risk and reference paths across endpoints (Fig. 3).

**Fig. 3.**
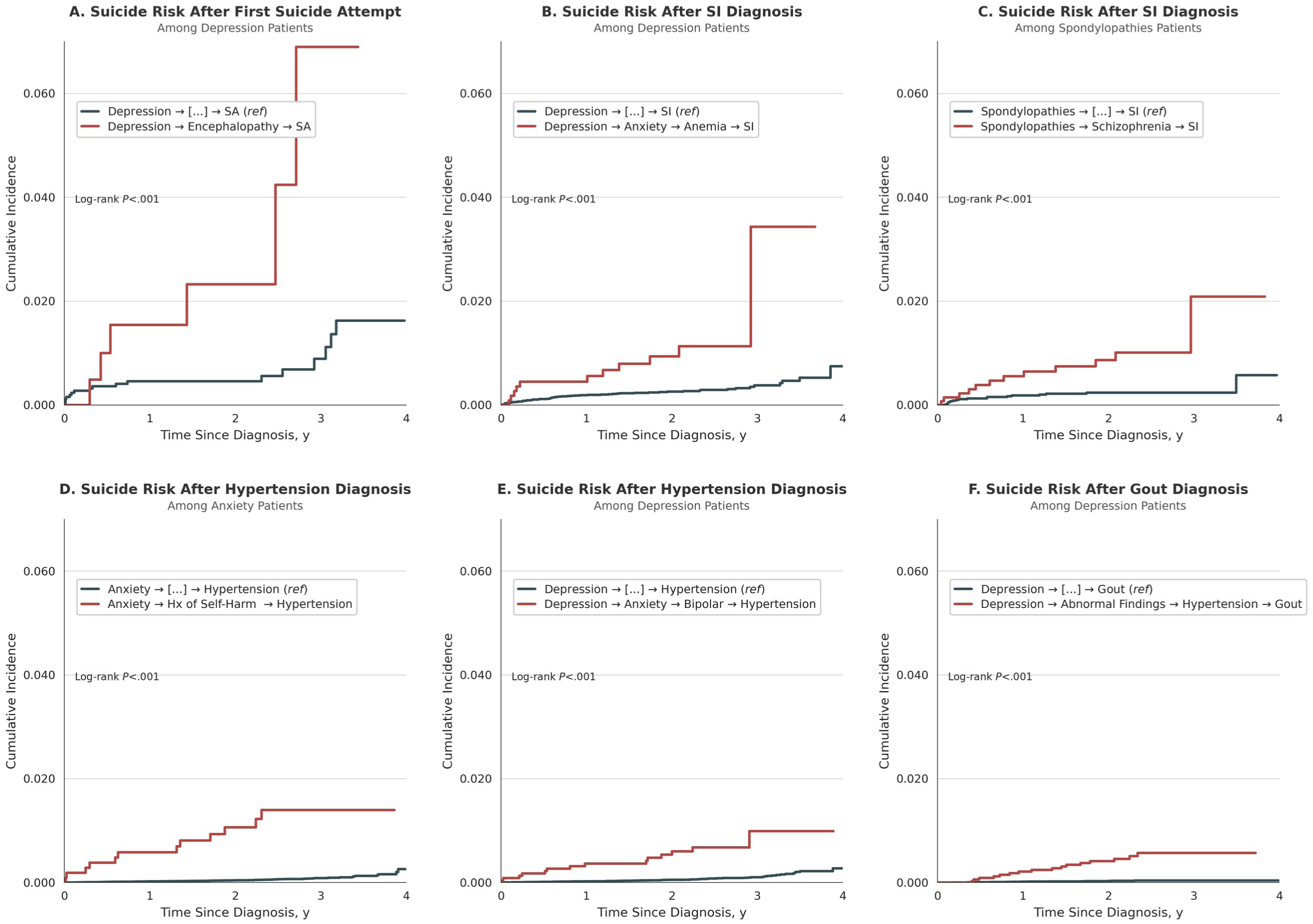
Cumulative incidence of suicide death after landmark diagnoses stratified by diagnostic path. Abbreviations: SA, suicide attempt; SI, suicidal ideation. Cumulative incidence of suicide death among patients sharing the same start and endpoint diagnoses, comparing those who followed a specific high-risk intermediate path (red) with those who reached the same endpoint through other intermediate diagnoses (black, reference). Time zero is the endpoint diagnosis date plus a lag period (30 days). Log-rank *P* values compare the high-risk path with the reference group.

**Table 1.** Suicide risk after landmark diagnoses by diagnostic path.

| Trajectory <sup>a</sup> | N <sup>b</sup> | Path <sup>c</sup> | Exposed, n (%) | Events | IR/1000 PY <sup>d</sup> | AHR (95% CI) <sup>e</sup> |
| --- | --- | --- | --- | --- | --- | --- |
| **Ending in Suicide Attempt** |  |  |  |  |  |  |
| Depression → [...] → SA | 3081 | Encephalopathy | 243 (7.9) | <11 | 17.3 | 6.2 (2.3-16.8) |
| Depression → [...] → SA | 3081 | Headache | 539 (17.5) | <11 | 10.1 | 4.1 (1.7-10.0) |
| Depression → [...] → SA | 3081 | Implant/device | 477 (15.5) | <11 | 9.6 | 3.0 (1.1-7.7) |
| **Ending in Suicidal Ideation** |  |  |  |  |  |  |
| Anxiety → [...] → SI | 20 710 | Anemia | 2475 (12.0) | 16 | 3.6 | 3.6 (1.9-6.9) |
| Depression → [...] → SI | 26 356 | Anemia | 2571 (9.8) | 15 | 3.2 | 3.1 (1.6-6.0) |
| Depression → [...] → SI | 26 356 | Anxiety | 11 155 (42.3) | 37 | 1.6 | 1.3 (0.8-2.1) |
| Depression → [...] → SI | 26 356 | Anxiety → Anemia | 1186 (4.5) | 11 | 5.2 | 4.6 (2.2-9.5) |
| Depression → [...] → SI | 26 356 | Alcohol use disorder | 4240 (16.1) | 15 | 1.8 | 1.2 (0.7-2.2) |
| Spondylopathies → [...] → SI | 10 001 | Schizophrenia | 1422 (14.2) | 12 | 4.4 | 4.5 (2.1-9.4) |
| **Ending in Hypertension** |  |  |  |  |  |  |
| Anxiety → [...] → HTN | 149 215 | Hx of self-harm | 1096 (0.7) | 12 | 5.0 | 32.0 (16.6-61.6) |
| Depression → [...] → HTN | 122 697 | Anxiety | 41 582 (33.9) | 39 | 0.4 | 1.5 (1.0-2.4) |
| Depression → [...] → HTN | 122 697 | Anxiety → Bipolar | 2466 (2.0) | 14 | 2.5 | 10.3 (5.6-19.1) |
| Depression → [...] → HTN | 122 697 | Anxiety → Hx of self-harm | 679 (0.6) | <11 | 5.4 | 20.7 (9.5-44.8) |
| **Ending in Metabolic Dysfunction** |  |  |  |  |  |  |
| Depression → [...] → Lipid disorders | 143 317 | HTN | 56 721 (39.6) | 45 | 0.3 | 1.2 (0.8-1.8) |
| Depression → [...] → Lipid disorders | 143 317 | Nausea/vomiting | 13 407 (9.4) | 24 | 0.8 | 3.7 (2.2-6.1) |
| Depression → [...] → Lipid disorders | 143 317 | Nausea/vomiting → HTN | 4741 (3.3) | 12 | 1.1 | 4.4 (2.3-8.4) |
| Depression → [...] → Gout | 51 269 | Abnormal findings → Fatigue | 3834 (7.5) | 16 | 2.0 | 8.5 (4.1-17.4) |
| Depression → [...] → Gout | 51 269 | Abnormal findings → HTN | 3627 (7.1) | 16 | 2.0 | 9.7 (4.2-22.8) |
| **Ending in Bipolar and Related Disorders** |  |  |  |  |  |  |
| Depression → [...] → Bipolar | 44 846 | Anxiety | 22 821 (50.9) | 34 | 0.7 | 1.6 (0.9-2.9) |
| Depression → [...] → Bipolar | 44 846 | Anxiety → Skin disorders | 3284 (7.3) | 11 | 1.7 | 3.7 (1.8-7.5) |
| Depression → [...] → Bipolar | 44 846 | Schizophrenia | 5017 (11.2) | 15 | 1.5 | 3.3 (1.8-5.9) |
| **Ending in SDOH** |  |  |  |  |  |  |
| Anxiety → [...] → SDOH | 35 272 | SI | 4561 (12.9) | 20 | 2.3 | 7.2 (3.7-13.9) |
| Depression → [...] → SDOH | 33 204 | SI | 5876 (17.7) | 23 | 2.0 | 6.4 (3.3-12.6) |
| General symptoms → [...] → SDOH | 28 529 | SI | 2426 (8.5) | 14 | 3.2 | 14.2 (6.1-32.9) |
| **Ending in Opioid-Related Disorders** |  |  |  |  |  |  |
| Depression → [...] → Opioid disorders | 18 836 | Musculoskeletal pain | 4915 (26.1) | 14 | 1.4 | 2.6 (1.1-6.0) |

Abbreviations: AHR, adjusted hazard ratio; CI, confidence interval; IR, incidence rate; PY, person-years; SA, suicide attempt; SI, suicidal ideation; HTN, hypertension; Hx, history; SDOH, social determinants of health.
Each row represents a specific path compared with all other patients completing the same start → end transition. Rows are non-mutually exclusive. AHRs are not comparable across trajectories. Event counts reflect post-landmark suicide deaths after the 30-day lag period and may be lower than the mining frequency threshold of 11 due to deaths occurring during the lag. Event counts fewer than 11 are reported as <11.
<sup>a</sup> Trajectory defines the start → end transition.
<sup>b</sup> Total patients completing the transition (landmark cohort).
<sup>c</sup> Specific intermediate diagnoses; reference = other paths with the same start → end trajectory (not shown).
<sup>d</sup> Incidence rate of suicide death per 1000 person-years.
<sup>e</sup> Adjusted for age, sex, race/ethnicity, strata (entry year), observable duration, 12-month coverage completeness, Charlson comorbidity index at trajectory start.

### Effect Modification by Sex and Age

Path-specific associations were generally consistent across sex and age groups (Table 2). Three paths showed significant sex interactions (*P* = 0.01, 0.007, and 0.008, respectively): among patients with depression who developed disorders of lipid metabolism, the nausea/vomiting → hypertension path was more strongly associated with suicide risk in women (AHR, 8.2; 95% CI, 3.8-17.7) than men (AHR, 1.4; 95% CI, 0.3-5.9); among patients with depression who reached SDOH through SI, women had higher risk (AHR, 22.4; 95% CI, 6.2-80.9) than men (AHR, 3.2; 95% CI, 1.4-7.3); and among patients with general symptoms who reached SDOH through SI, women similarly had higher risk (AHR, 91.4; 95% CI, 11.3-741.8) than men (AHR, 6.4; 95% CI, 2.2-18.3). The anemia path from anxiety to SI showed an age interaction (*P* = 0.04), with stronger association among adults aged 44 years or younger (AHR, 6.8; 95% CI, 2.6-17.5) than those aged 65 years or older (AHR, 0.6; 95% CI, 0.1-4.7).

**Table 2.** Path-specific suicide risk by sex and age.

| Trajectory <sup>a</sup> | Path | Sex, AHR (95% CI) |  | Age, AHR (95% CI), y |  |  |
| --- | --- | --- | --- | --- | --- | --- |
|  |  | Female | Male | ≤44 | 45-64 | ≥65 |
| Ending in Suicide Attempt |  |  |  |  |  |  |
| Depression → [...] → SA | Encephalopathy | 9.4 (3.0-29.1) | 2.2 (0.3-18.3) | 4.1 (0.9-18.6) | 13.9 (3.1-62.2) | † |
| Depression → [...] → SA | Headache | 6.5 (2.3-18.7) | 1.1 (0.1-9.1) | 4.5 (1.6-13.3) | 4.5 (1.0-19.5) | † |
| Depression → [...] → SA | Implant/device | 4.3 (1.5-12.6) | 1.0 (0.1-8.4) | 3.8 (1.2-12.7) | 3.3 (0.8-13.8) | † |
| Ending in Suicidal Ideation |  |  |  |  |  |  |
| Anxiety → [...] → SI | Anemia <sup>b</sup> | 3.7 (1.6-8.6) | 3.5 (1.4-8.5) | 6.8 (2.6-17.5) | 4.5 (1.9-10.6) | 0.6 (0.1-4.7) |
| Depression → [...] → SI | Anemia | 3.2 (1.4-7.4) | 2.9 (1.1-7.4) | 5.3 (1.9-14.3) | 4.2 (1.8-9.7) | 0.6 (0.1-4.6) |
| Depression → [...] → SI | Anxiety → Anemia | 4.1 (1.6-10.5) | 5.4 (2.0-14.6) | 11.2 (4.1-30.7) | 4.3 (1.6-11.8) | 1.3 (0.2-10.8) |
| Spondylopathies → [...] → SI | Schizophrenia | 3.4 (1.2-10.0) | 5.8 (2.1-16.2) | 6.1 (1.6-22.8) | 5.0 (1.8-14.2) | 2.7 (0.5-14.8) |
| Ending in Hypertension |  |  |  |  |  |  |
| Anxiety → [...] → HTN | Hx of self-harm | 36.2 (14.4-90.5) | 28.8 (11.9-69.6) | 22.9 (9.1-57.6) | 38.2 (13.4-109.2) | 51.3 (11.7-224.6) |
| Depression → [...] → HTN | Anxiety → Bipolar | 13.8 (6.1-31.3) | 7.8 (3.2-18.8) | 8.0 (3.4-19.1) | 12.6 (4.8-32.9) | 12.2 (2.8-52.6) |
| Depression → [...] → HTN | Anxiety → Hx of self-harm | 24.0 (8.2-70.5) | 18.2 (6.4-52.0) | 12.5 (4.3-36.1) | 36.1 (10.9-119.0) | 35.6 (4.8-266.8) |
| Ending in Metabolic Dysfunction |  |  |  |  |  |  |
| Depression → [...] → Lipid disorders | Nausea/vomiting | 4.5 (2.2-9.1) | 3.2 (1.6-6.2) | 4.9 (2.2-10.7) | 4.0 (1.9-8.3) | 1.9 (0.6-6.4) |
| Depression → [...] → Lipid disorders | Nausea/vomiting → HTN <sup>c</sup> | 8.2 (3.8-17.7) | 1.4 (0.3-5.9) | 4.2 (1.7-10.8) | 6.9 (2.8-16.8) | † |
| Depression → [...] → Gout | Abnormal findings → Fatigue | 10.6 (3.3-33.9) | 7.1 (2.5-19.9) | 6.1 (1.7-22.4) | 14.7 (3.5-61.6) | 7.5 (2.2-26.4) |
| Depression → [...] → Gout | Abnormal findings → HTN | 21.1 (5.4-82.2) | 5.7 (1.9-17.1) | 19.3 (3.9-96.0) | 16.7 (3.9-71.3) | 2.9 (0.6-14.4) |
| Ending in Bipolar and Related Disorders |  |  |  |  |  |  |
| Depression → [...] → Bipolar | Anxiety → Skin disorders | 3.8 (1.5-9.7) | 3.5 (1.3-9.6) | 6.2 (2.4-16.0) | 3.1 (1.0-9.5) | 1.2 (0.2-9.8) |
| Depression → [...] → Bipolar | Schizophrenia | 4.5 (1.9-10.3) | 2.5 (1.0-5.7) | 2.6 (1.0-6.4) | 3.4 (1.2-9.3) | 5.8 (1.5-21.7) |
| Ending in SDOH |  |  |  |  |  |  |
| Anxiety → [...] → SDOH | SI | 10.4 (4.0-27.0) | 5.3 (2.2-12.7) | 3.8 (1.5-9.9) | 9.1 (3.4-24.6) | 22.6 (4.1-124.5) |
| Depression → [...] → SDOH | SI <sup>c</sup> | 22.4 (6.2-80.9) | 3.2 (1.4-7.3) | 3.0 (1.1-8.1) | 10.9 (3.7-32.2) | 9.5 (2.1-42.8) |
| General symptoms → [...] → SDOH | SI <sup>c</sup> | 91.4 (11.3-741.8) | 6.4 (2.2-18.3) | 4.0 (1.0-16.9) | 31.9 (8.1-124.6) | 25.6 (4.6-142.8) |
| Ending in Opioid-Related Disorders |  |  |  |  |  |  |
| Depression → [...] → Opioid disorders | Musculoskeletal pain | 2.7 (0.7-10.0) | 2.5 (0.9-6.9) | 4.5 (1.5-13.1) | 1.5 (0.4-5.9) | 0.9 (0.1-10.3) |

Abbreviations: AHR, adjusted hazard ratio; CI, confidence interval; SA, suicide attempt; SI, suicidal ideation; Hx, history; HTN, hypertension; SDOH, social determinants of health.
<sup>a</sup> Each path compared with all other patients completing the same start → end transition (binary comparison, same design as Table 1). Adjusted for age, sex, race/ethnicity, strata (entry year), observable duration, 12-month coverage completeness, Charlson comorbidity index at trajectory start. <sup>b</sup> Significant age × path interaction ( $P < 0.05$ ; likelihood ratio test). <sup>c</sup> Significant sex × path interaction ( $P < 0.05$ ; likelihood ratio test).
† Insufficient events for estimation.

### Progressive Adjustment

For trajectories with sufficient events to define mutually exclusive paths, hazard ratios were stable across progressively adjusted models (Table 3). For example, the anxiety → bipolar path to hypertension had an AHR of 12.1 (95% CI, 6.5-22.4) in the demographic model and 10.9 (95% CI, 5.7-20.8) after full adjustment. Notably, path-specific estimates remained elevated after adjustment for comorbidity burden, whether measured at trajectory start or at endpoint. Hazard ratios were additionally stable after adjustment for trajectory duration (Additional file 1: Table S5).

**Table 3.** Path-stratified suicide risk within diagnostic trajectories.

| Trajectory <sup>a</sup> | Path | N | Events | IR/1000 PY | Model, AHR (95% CI) <sup>b</sup> |  |  |
| --- | --- | --- | --- | --- | --- | --- | --- |
|  |  |  |  |  | Model 1 | Model 2 | Model 3 |
| Depression → [...] → SI | Other paths | 15 708 | 36 | 1.1 | 1 [Reference] | 1 [Reference] | 1 [Reference] |
|  | Anxiety | 9462 | 25 | 1.3 | 1.1 (0.7-1.9) | 1.1 (0.6-1.8) | 1.1 (0.6-1.8) |
|  | Anxiety → Anemia | 1186 | 11 | 5.2 | 4.8 (2.4-9.8) | 4.8 (2.2-10.5) | 7.8 (3.5-17.3) |
| Depression → [...] → Hypertension | Other paths | 81 760 | 51 | 0.2 | 1 [Reference] | 1 [Reference] | 1 [Reference] |
|  | Anxiety | 38 471 | 25 | 0.3 | 1.2 (0.7-1.9) | 1.1 (0.7-1.8) | 1.1 (0.7-1.9) |
|  | Anxiety → Bipolar | 2466 | 14 | 2.5 | 12.1 (6.5-22.4) | 10.9 (5.7-20.8) | 10.9 (5.7-20.9) |
| Depression → [...] → Hypertension | Other paths | 81 249 | 51 | 0.2 | 1 [Reference] | 1 [Reference] | 1 [Reference] |
|  | Anxiety | 40 769 | 31 | 0.3 | 1.4 (0.9-2.2) | 1.3 (0.8-2.1) | 1.3 (0.8-2.1) |
|  | Anxiety → Hx of self-harm | 679 | <11 | 5.4 | 26.9 (12.2-59.2) | 23.3 (10.4-51.9) | 23.5 (10.5-52.6) |
| Depression → [...] → Bipolar | Other paths | 23 990 | 23 | 0.4 | 1 [Reference] | 1 [Reference] | 1 [Reference] |
|  | Anxiety | 17 572 | 22 | 0.6 | 1.4 (0.8-2.5) | 1.4 (0.8-2.6) | 1.5 (0.8-2.7) |
|  | Anxiety → Skin disorders | 3284 | 11 | 1.7 | 4.5 (2.2-9.4) | 4.3 (2.0-9.4) | 5.2 (2.4-11.2) |
| Depression → [...] → Lipid disorders | Other paths | 82 846 | 45 | 0.2 | 1 [Reference] | 1 [Reference] | 1 [Reference] |
|  | Hypertension | 49 522 | 29 | 0.2 | 1.0 (0.6-1.6) | 0.9 (0.6-1.5) | 0.9 (0.6-1.4) |
|  | Nausea/vomiting | 6208 | <11 | 0.6 | 3.2 (1.5-6.8) | 2.7 (1.2-5.8) | 2.6 (1.2-5.7) |
|  | Nausea/vomiting → HTN | 4741 | 12 | 1.1 | 6.3 (3.2-12.2) | 4.7 (2.3-9.3) | 4.4 (2.2-8.8) |
| Anxiety → [...] → SDOH | Other paths | 31 087 | 19 | 0.3 | 1 [Reference] | 1 [Reference] | 1 [Reference] |
|  | SI | 3267 | 14 | 2.2 | 6.3 (3.1-12.8) | 6.5 (3.2-13.4) | 6.5 (3.2-13.2) |
|  | General symptoms → SI | 918 | <11 | 3.8 | 10.7 (4.2-27.1) | 13.4 (5.1-35.6) | 15.0 (5.6-39.9) |
| Depression → [...] → SDOH | Other paths | 27 823 | 16 | 0.3 | 1 [Reference] | 1 [Reference] | 1 [Reference] |
|  | SI | 4441 | 16 | 1.8 | 5.5 (2.7-11.5) | 5.3 (2.6-11.1) | 5.3 (2.6-11.0) |
|  | General symptoms → SI | 940 | <11 | 3.6 | 10.8 (4.2-27.8) | 10.5 (3.9-27.7) | 12.3 (4.6-32.8) |

Abbreviations: AHR, adjusted hazard ratio; CI, confidence interval; IR, incidence rate; PY, person-years; SI, suicidal ideation; Hx, history; HTN, hypertension; SDOH, social determinants of health.
<sup>a</sup> Trajectories with sufficient events across multiple intermediate paths were further stratified to enable direct path comparison. Within each block, patients who completed the same start → end transition were stratified into mutually exclusive paths. The reference group includes patients who reached the endpoint through paths not explicitly modeled.
<sup>b</sup> Model 1: age, sex, race/ethnicity, strata (entry year). Model 2 (primary): Model 1 + observable duration, 12-month coverage completeness, Charlson comorbidity index at trajectory start. Model 3 (sensitivity): Model 2 with Charlson comorbidity index at trajectory end.

## Discussion

In this retrospective cohort study of 3.6 million insured individuals in Maryland, we identified 1 816 diagnostic trajectories associated with suicide death. Longer sequences were more common among suicide decedents and generally carried greater risk than single conditions. Among individuals with the same origin and landmark diagnosis, post-landmark suicide risk varied according to the intermediate conditions accumulated over time. Prior work has shown that suicide decedents comprise clinically distinct subtypes [20,21], and that the ordering of mental disorders and medical conditions influences suicide risk [22]. Our findings extend this work by showing that intermediate diagnoses further distinguish risk.

Several high-risk trajectories align with clinical patterns familiar to psychiatrists. Depression followed by psychosis and then SI reflects a particularly high-risk phenotype often recognized in practice [8,23]. Trajectories from depression to alcohol-related disorders or chronic pain suggest how psychiatric symptoms, self-medication, and physical illness may reinforce one another [22,24–26]. Progression from depression or anxiety to ideation or attempts after prolonged psychotropic treatment may mark more persistent or treatment-complex illness.

Beyond these recognizable patterns, less obvious risk signals emerged spanning medical and psychiatric settings. A central finding was the role of physical conditions within psychiatric trajectories. Paths incorporating iron deficiency anemia, encephalopathy, headache, spondylopathies, skin disorders, nausea and vomiting, fatigue and malaise, or abnormal findings without diagnosis consistently conferred greater risk than more direct psychiatric paths to the same endpoint. This is consistent with literature linking a broad range of physical conditions to elevated suicide risk, including inflammation, metabolic dysfunction, and neurologic symptoms [27–30]. Encephalopathy preceding SA could reflect neurotoxicity, sequelae of prior overdose, or metabolic dysfunction affecting cognition and impulse control [31–33]. The headache to SA trajectory may reflect shared serotonergic mechanisms, chronic pain burden, or both [34,35]. Similarly, among patients with depression who developed SI, anemia-containing paths conferred substantially higher risk than paths through anxiety alone, suggesting that SI is a heterogeneous clinical state and that the diagnostic history preceding it carries prognostic information. Anemia may reflect nutritional deficiency, chronic inflammation, or medication side effects, each associated with depression severity and treatment resistance [36–38]. Spondylopathies marked higher-risk courses when appearing in trajectories ending in SA or SI, or at the origin of psychosis-spectrum trajectories.

Among cardiometabolic diagnoses, hypertension appeared most often, occurring in 10% of significant trajectories and occupying both intermediate and endpoint positions. Across trajectories involving hypertension, history of self-harm was the strongest intermediate diagnosis for separating subsequent suicide risk. Hypertension trajectories identified largely non-overlapping patient groups (Additional file 1: Fig. S6), suggesting different morbidity patterns. For example, the depression → anxiety → bipolar → hypertension trajectory may reflect diagnostic revision from unipolar depression to bipolar disorder [8,33,39]. By contrast, the trajectory depression → nausea/vomiting → hypertension → disorders of lipid metabolism, which was stronger in women, may involve psychiatric medication-related blood pressure crises [39]. In the longest metabolic cascades, hypertension linked depression through abnormal clinical findings to gout, consistent with accumulating metabolic and inflammatory burden [40–43]. These findings support the potential value of hypertension as a nonpsychiatric landmark for risk stratification.

For trajectories ending in SDOH, SI as an intermediate condition was associated with elevated risk. However, the highest association was observed among SI patients whose trajectory began with nonspecific general signs and symptoms rather than a psychiatric diagnosis. The general symptoms category in this pathway was dominated by ICD codes for edema, unexplained weight changes, unspecified pain, and lymphadenopathy, suggesting that somatization of psychological distress or undiagnosed physical illness may mark a particularly high-risk trajectory toward psychosocial adversity. Somatic complaints also appeared in the substance use domain, where trajectories such as depression → musculoskeletal pain → opioid-related disorders may capture a clinical progression in which undertreated or severe pain in patients with depression leads to opioid exposure and subsequent substance use disorder.

Most associations were consistent across sex and age and remained stable after progressive adjustment, suggesting they were not explained by baseline illness severity or comorbidity accumulated along the trajectory. Nonetheless, three paths showed sex interactions, each with stronger associations in women: one involving a cardiometabolic trajectory through nausea/vomiting and hypertension, and two involving SDOH trajectories through SI from different origins. The anemia path from anxiety to SI also showed an age interaction, with stronger association among younger adults. This suggests that some of these trajectories may carry sex- or age-dependent risk and warrant further investigation in different cohorts.

The prolonged duration of these trajectories and the extended interval between trajectory completion and death indicate that risk often accumulated over months to years, with a potential window for detection well before the terminal event. Suicide risk rarely appeared in isolated psychiatric encounters; more than half of decedents’ trajectories spanned multiple specialties, and such cross-domain signals may not be visible to individual providers when care for the same patient occurs across separate health systems. When patients present with SI or SA, the preceding diagnostic history may inform clinical assessment, particularly when physical conditions or nonspecific somatic findings mark intermediate stages of risk accumulation. The elevated risk observed at routine medical landmarks, including hypertension, lipid disorders, and gout, further suggests that opportunities for risk detection may exist in general medical care settings. Together, these findings emphasize the value of longitudinal, system-level surveillance approaches for suicide risk identification that can detect fragmented or evolving trajectories across the continuum of care [44,45].

### Limitations

This study has limitations. The cohort comprised insured individuals in Maryland and did not include Medicaid beneficiaries, who may have different clinical trajectories and utilization patterns, limiting generalizability. However, the statewide claims-based design, linkage to medical examiner records, large sample size, and population-level coverage are important strengths. We made several analytic choices to address potential sources of bias, as described in the Methods, but residual confounding cannot be excluded because claims data lack information on symptom severity, treatment response, and other unmeasured clinical factors. In addition, 38% of suicide decedents were not exposed to any significant trajectory, likely reflecting limited clinical engagement or surveillance gaps (Additional file 1: Table S4). The order of recorded diagnoses reflects clinical documentation rather than disease onset; observed trajectories may therefore reflect true clinical progression, shared underlying vulnerability, or documentation patterns. Nonetheless, associations were consistent and interpretable across multiple clinical domains, suggesting that diagnostic order may carry both prognostic and phenotypic signal. These findings should be viewed as hypothesis generating, and replication in independent populations is needed.

### Conclusions

We identified ordered diagnostic trajectories from statewide claims data that were associated with suicide death. Medical conditions and nonspecific somatic findings within psychiatric trajectories identified higher-risk clinical courses. Elevated suicide risk was also observed after sequences ending in routine medical conditions. These findings suggest that the temporal accumulation and order of clinical conditions carry prognostic value for suicide, that suicide risk signals may be fragmented across clinical settings, and that opportunities for risk detection may extend beyond traditional psychiatric settings.

## List of abbreviations

AHR: adjusted hazard ratio
CCSR: Clinical Classifications Software Refined
CI: confidence interval
CRISP: Chesapeake Regional Information System for Patients
FAC: factors influencing health status
HTN: hypertension
Hx: history
ICD-10-CM: International Classification of Diseases, Tenth Revision, Clinical Modification
IQR: interquartile range
IR: incidence rate
LR: logistic regression
MSDW: Maryland Suicide Data Warehouse
OCME: Office of the Chief Medical Examiner
PY: person-years
SA: suicide attempt
SD: standard deviation
SDOH: social determinants of health
SI: suicidal ideation
STROBE: Strengthening the Reporting of Observational Studies in Epidemiology.

## Declarations

### Ethics approval and consent to participate

This retrospective cohort study was approved by the Johns Hopkins Bloomberg School of Public Health Institutional Review Board (#00009022) with a waiver of informed consent.

### Consent for publication

Not applicable.

### Availability of data and materials

The datasets analyzed during this study contain protected health information subject to the Health Insurance Portability and Accountability Act and cannot be publicly shared. Access may be granted upon reasonable request to the corresponding author, subject to permission from data providers in the Maryland Suicide Data Warehouse and institutional review board approval. Analysis code is available from the corresponding author upon reasonable request.

### Authors’ contributions

A.B., C.K., and H.K. had full access to all the data in the study and take responsibility for the integrity of the data and the accuracy of the data analysis. Concept and design: A.B., C.K., E.H., H.L., P.S.N., H.C.W., H.K. Acquisition, analysis, or interpretation of data: A.B., C.K., E.H., H.L., P.S.N., H.C.W., H.K. Drafting of the manuscript: A.B. Critical revision of the manuscript for important intellectual content: All authors. Statistical analysis: A.B., C.K. Obtained funding: E.H., H.K., H.C.W., P.S.N. Administrative, technical, or material support: C.K., H.K. Supervision: H.K., P.S.N., H.C.W. All authors read and approved the final manuscript.

## Acknowledgements

The authors thank the Maryland Health Care Commission, the Maryland Office of the Chief Medical Examiner, and the Chesapeake Regional Information System for Patients (CRISP) for providing access to the data used in this study.

## Additional files

Additional file 1 (.docx): Supplementary material. Supplementary Methods, Figs. S1–S6, and Tables S1–S5.

## Supplementary Methods

### CCSR version selection

Given the cross-setting and multiyear study period (January 2016 through December 2020), we required a classification system that provides consistent categorization across care settings and accounts for ICD-10-CM code updates during this timeframe. We selected the Clinical Classifications Software Refined (CCSR) framework, version 2025.1. This version was chosen over the earlier v2021.1 after comparison: the 2025 version contains 2 033 additional codes not present in the 2021 version while maintaining all earlier codes. It also introduced clinically significant distinctions, including specific categories for conditions in remission (previously grouped with active disease), refinements to injury classification, and improved handling of codes classified differently between inpatient and outpatient settings, ensuring consistency in cross-setting analyses.

### Suicide-related code reclassification

Suicide-related ICD-10-CM codes are dispersed across numerous CCSR categories spanning mental, behavioral, and neurodevelopmental disorders; injury; external causes; factors influencing health status; and symptoms and signs. This dispersion obscures important clinical distinctions for suicide research. To address this, we adopted the CDC/NCHS surveillance case definitions for suicide attempt and intentional self-harm, reclassified the relevant ICD-10-CM codes within the CCSR framework, and introduced distinctions between suicidal ideation, suicide attempts, and self-harm history.

The reclassification resulted in three self-harm categories (SH001, SH002, SH003), two suicide attempt categories (SA001, SA002), and one suicidal ideation category (SI). Poisoning codes were mapped by intent: intentional self-harm to SH001, undetermined intent to SH003, while accidental, assault, adverse effect, and underdosing codes remained in their original CCSR categories. This yielded 1 040 codes in SH001, 78 in SH002, 3 in SA001, 3 in SA002, and 1 in SI. Several granular self-harm codes (R45.88, Z91.51, Z91.52) introduced in October 2021 did not appear in our 2016-2020 dataset; however, the reclassification was applied to the complete CCSR reference file to ensure future applicability. The full mapping is provided in **Table S1**.

### Reclassification of factors influencing health status (chapter 21) codes

Chapter 21 of ICD-10-CM (Factors influencing health status and contact with health services) combines true health states (eg, disability, history of cancer) with administrative encounters (eg, routine examinations, immunizations). To reduce detection bias and avoid treating administrative encounters as clinical risk states, we manually reclassified these codes.

We excluded codes representing purely administrative or preventive contact, including routine medical examinations, screening-only encounters, immunizations, contraceptive management, generic aftercare, and counseling not related to mental health. We retained codes capturing psychosocial adversity (eg, abuse-related mental health encounters, socioeconomic and lifestyle problems, tobacco use); chronic disease burden and disability (eg, dialysis, respiratory support, mobility or ostomy status, organ transplant, acquired organ or limb loss); treatment history and nonadherence (eg, personal history of self-harm, patient or caregiver noncompliance); and high-acuity care states (eg, physical restraint, need for supervision, hazardous exposures, do-not-resuscitate status).

Several broad CCSR categories that combined clinically heterogeneous conditions under a single code (eg, other specified status, long-term medication use, dependency or assistance status, body mass index) were further divided into clinically coherent subgroups (eg, dialysis, respiratory support, mobility dependence, long-term use of antithrombotic, diabetes, steroid, pain, or opioid therapy, underweight or overweight body mass index). Subgroups were retained when they reflected chronic illness, biologically plausible mechanisms, or elevated suicide counts. Very low-yield or purely administrative subcodes (eg, generic carrier status, organ donor status, family history codes) were excluded. The complete mapping of original codes to refined subgroups with inclusion and exclusion decisions is provided in **Table S2**.

### Sequential pattern mining with PrefixSpan algorithm

PrefixSpan (Prefix-Projected Sequential Pattern Mining) discovers frequent sequential patterns in ordered event data by taking each common starting event (a prefix) and projecting the database to what follows, extending a path only when every shorter prefix is itself frequent. Frequency is measured by support, defined as the proportion of patients whose histories contain the sequence at least once in that order. For example, if depression is frequent, PrefixSpan examines what commonly follows; if depression → anxiety also meets the support threshold, it is extended; if depression → anxiety → SI meets the threshold, it is retained; otherwise, growth stops. By growing only from frequent prefixes, the algorithm avoids brute-force enumeration of all possible orderings and yields a concise set of reproducible trajectories suitable for downstream association testing. The pseudocode for the PrefixSpan algorithm is shown in Algorithm 1.

**Algorithm 1** PrefixSpan Algorithm for Sequential Pattern Mining

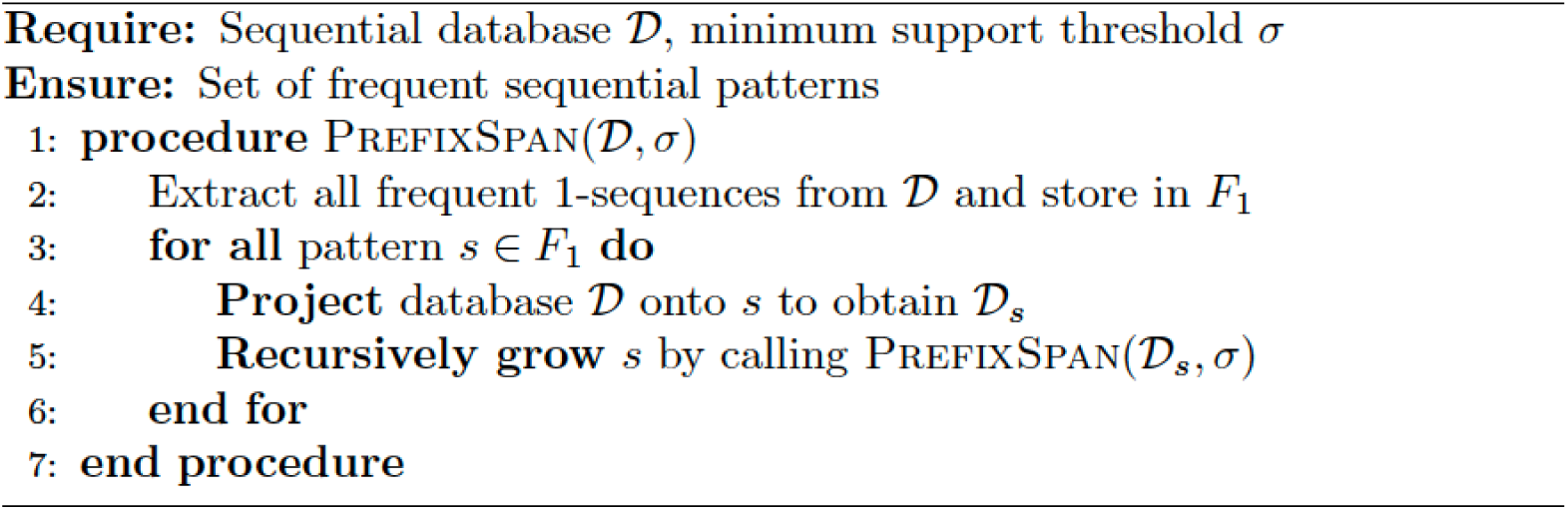

**Fig. S1.**
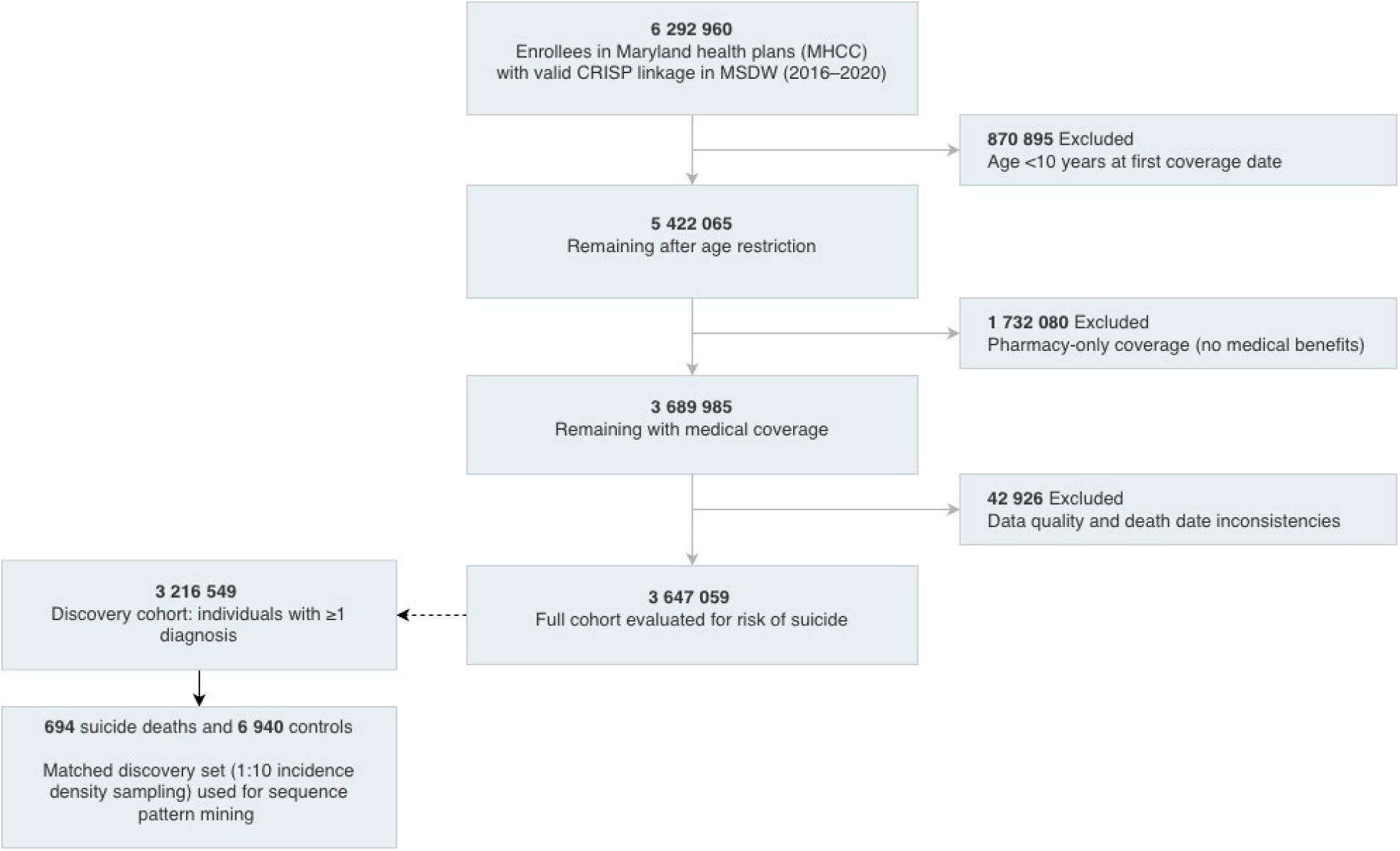
Study population flowchart. Abbreviations: MHCC, Maryland Health Care Commission; MSDW, Maryland Suicide Data Warehouse; CRISP, Chesapeake Regional Information System; OCME, Office of the Chief Medical Examiner. Note: Suicide deaths ascertained via OCME records.

**Fig. S2.**
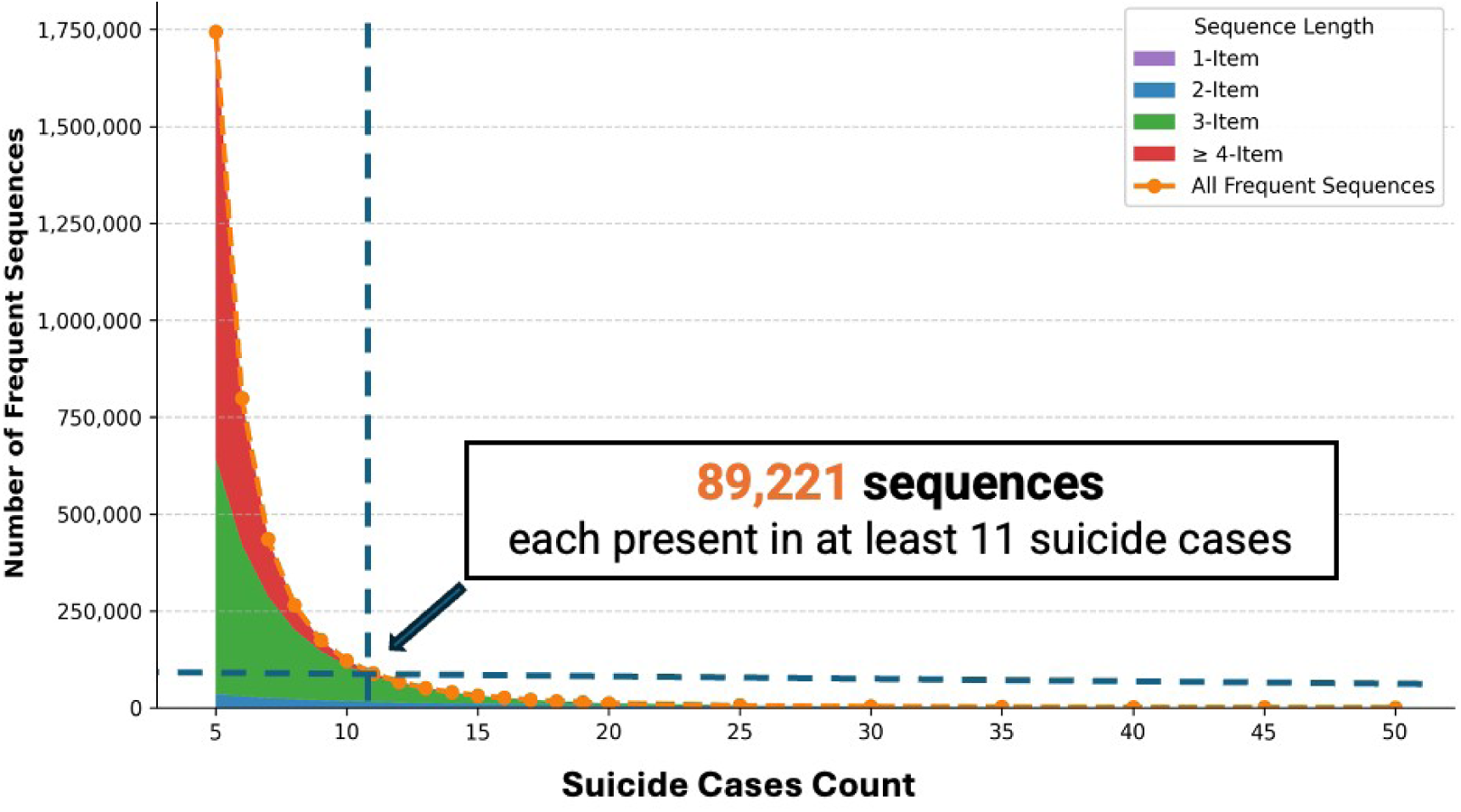
Number of candidate sequences by minimum frequency threshold. Threshold frequency distribution of 89 221 sequences meeting minimum frequency (≥11 suicide cases). Colors indicate sequence length (1- to 4-item). Most trajectories appear in few cases, with progressively fewer sequences appearing at higher frequencies.

**Fig. S3.**
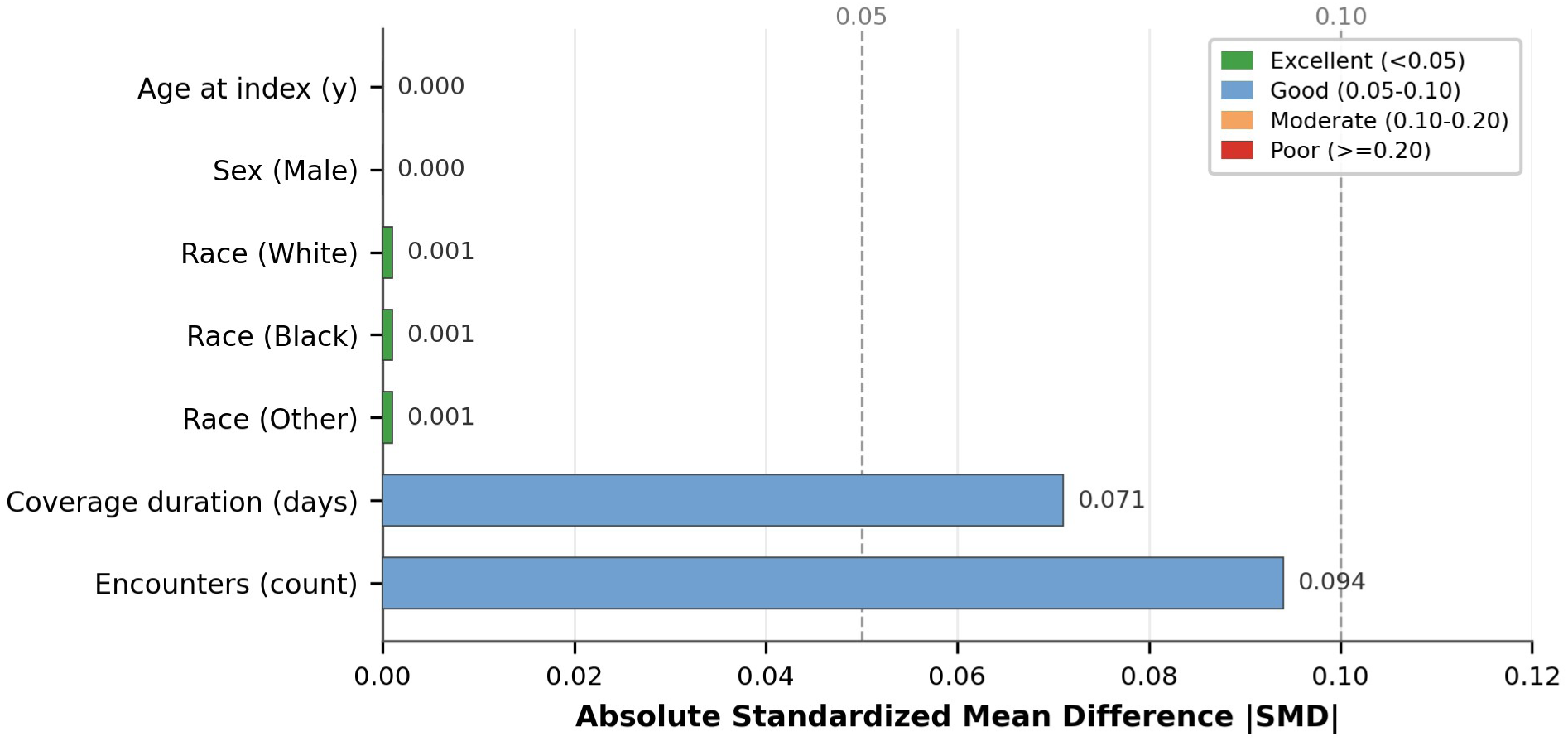
Covariate balance in the discovery cohort after incidence-density matching. Standardized mean differences (SMDs) for matching variables and related cohort characteristics in the matched discovery cohort. All covariates achieved acceptable balance (SMD < 0.10).

**Fig. S4.**
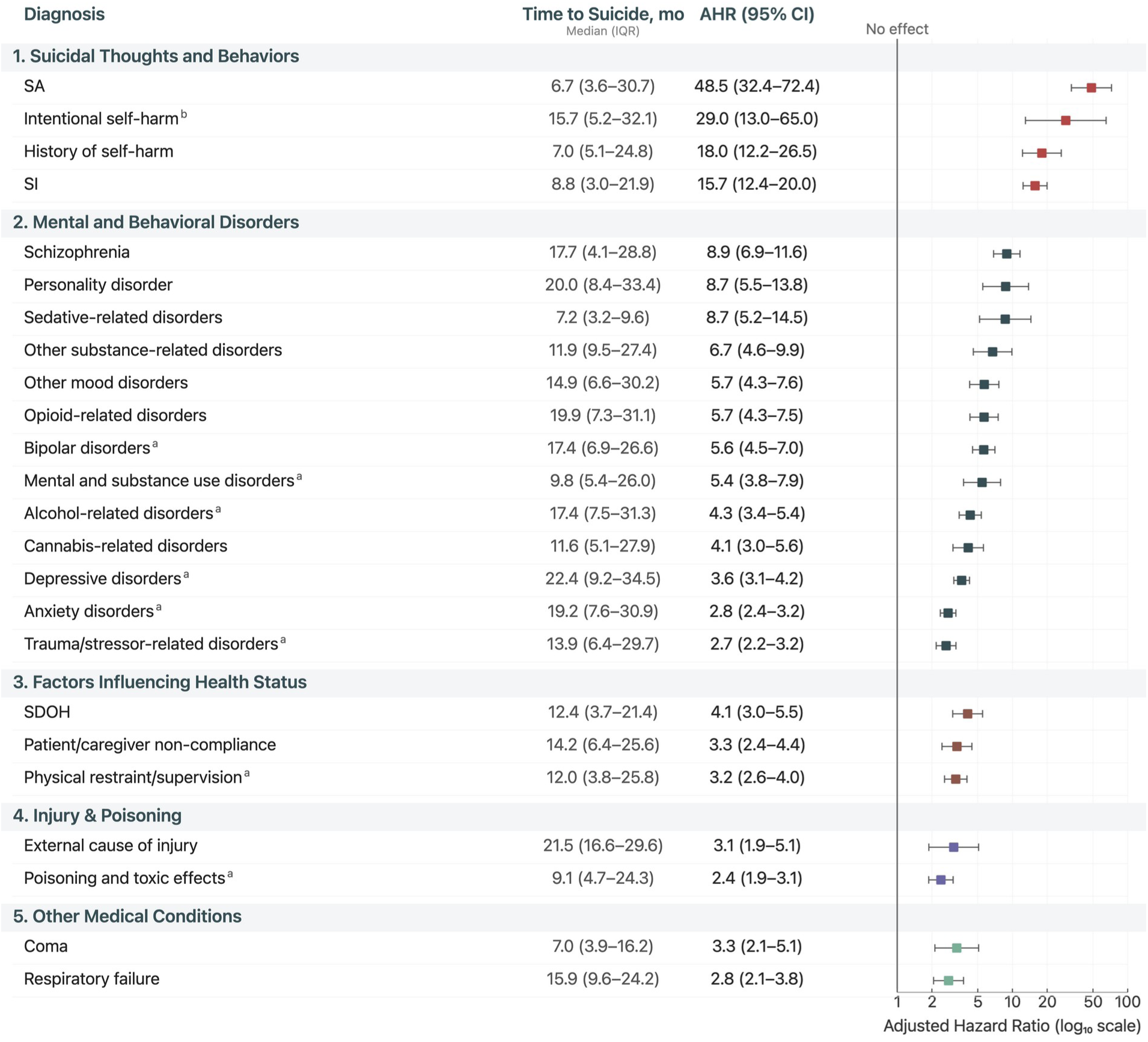
Individual conditions associated with suicide death. Abbreviations: SDOH, social determinants of health; SA, suicide attempt; SI, suicidal ideation. Of 1 816 significant sequences identified in time-varying Cox models, 24 were single-condition associations (Bonferroni-corrected P < 0.05). Multistep trajectories (≥2 conditions) are presented in Fig. 2 of the main manuscript alongside domain-specific single-condition baselines. Time to Suicide represents median time (IQR) from first diagnosis to suicide death among decedents. AHRs adjusted for age, sex, race/ethnicity, and cohort entry year. ^a^ Proportional hazards assumption violated; AHR represents time-averaged effect over follow-up.^b^ Fewer than 11 events after 30-day lag exclusion; unable to form sequential patterns in trajectory analysis.

**Fig. S5.**
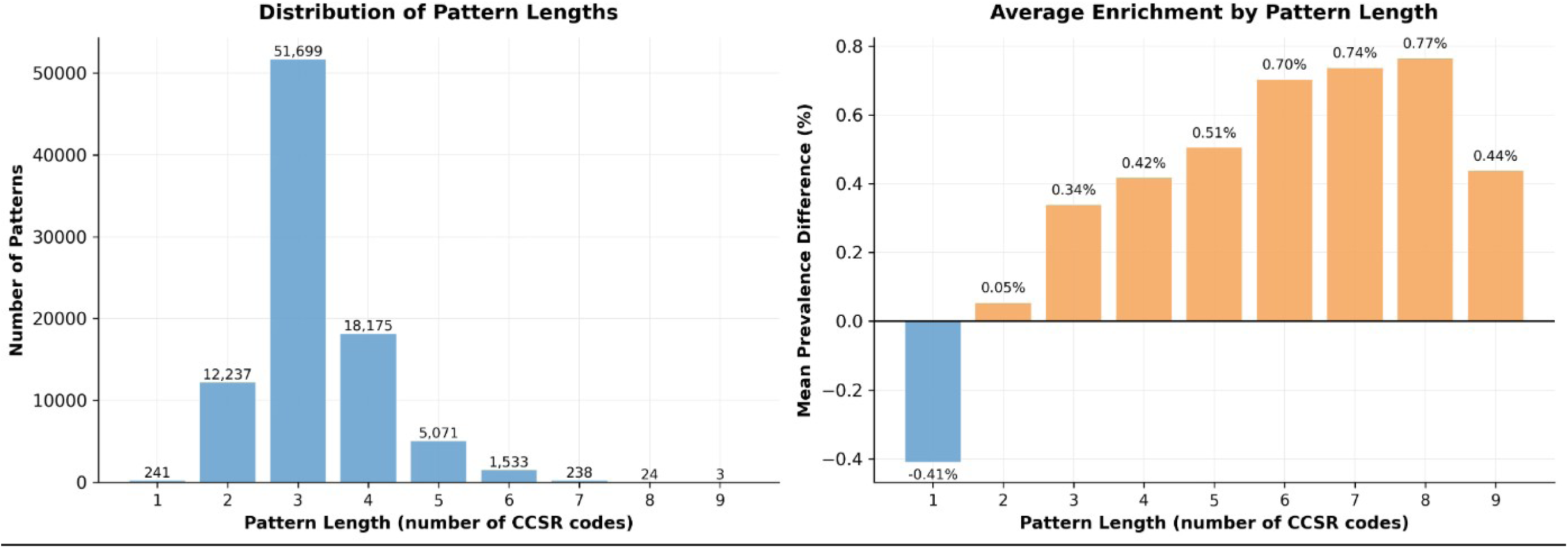
Distribution of candidate sequence lengths and case enrichment by length. Left: Number of mined sequences by length (number of CCSR codes). Right: Mean prevalence difference between suicide decedents and matched controls for sequences of each length. Single-code patterns are slightly more common in controls, whereas longer sequences show increasing enrichment among decedents.

**Fig. S6.**
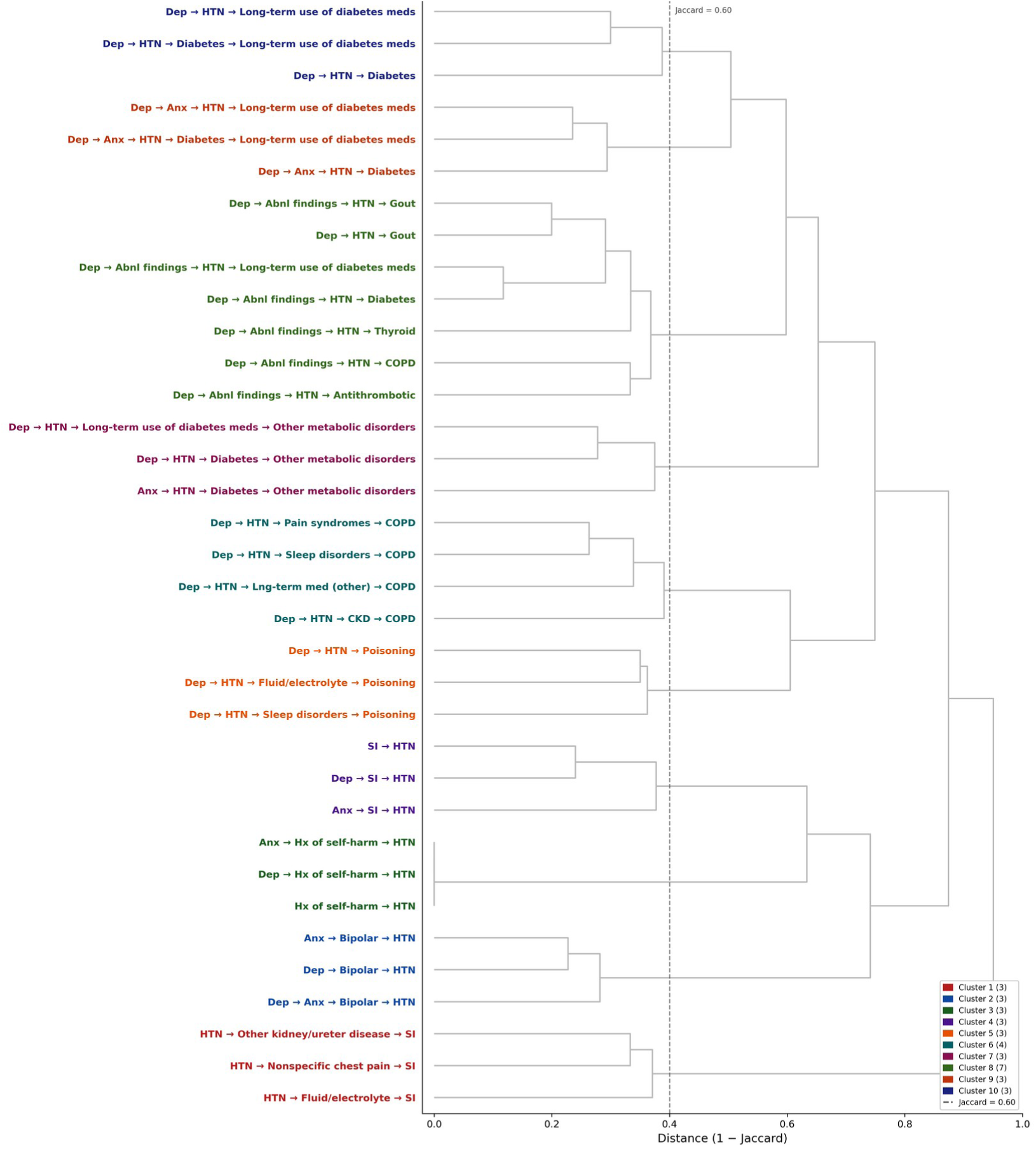
Patient overlap clustering of hypertension trajectories among suicide decedents. Abbreviations: Abnl, abnormal; Anx, anxiety; CKD, chronic kidney disease; COPD, chronic obstructive pulmonary disease; Dep, depression; HTN, hypertension; Hx, history; SI, suicidal ideation. Hierarchical clustering of 183 Bonferroni-significant hypertension-containing trajectories among suicide decedents, based on pairwise patient overlap quantified using the Jaccard similarity coefficient and displayed as distance (1 − Jaccard). Trajectories were clustered using average linkage, and the dashed vertical line indicates a Jaccard similarity threshold of 0.60 (distance = 0.40). To focus on reproducible groupings, only trajectories belonging to clusters containing at least 3 pathways at this threshold are displayed (35 of 183 trajectories); the remaining 148 trajectories showed limited overlap with other pathways and are not shown. Colored labels denote cluster membership. The displayed clusters suggest that hypertension-associated suicide risk is distributed across several partially distinct trajectory groupings, including bipolar-related, self-harm, suicidal ideation, metabolic, and somatic trajectories.

**Table S1.** Reclassification of suicide/self-harm ICD-10-CM codes.

| Category | Description | ICD-10-CM Codes | Clinical Relevance |
| --- | --- | --- | --- |
| SI | Suicidal ideation | R45.851 | Active thoughts of suicide without action |
| SA001 | Suicide attempt (acute) | T14.91, T14.91XA | Acute suicide attempt |
| SA002 | Personal history of suicidal behavior | T14.91XD, T14.91XS, Z91.51 | Post-attempt care, follow-up, or historical suicidal behavior |
| SH001 | Intentional self-harm | X60-X84; T36-T65 with 6th character "2"; T71 with 6th character "2"; Y87.0; R45.88 | Intentional self-injury or poisoning (suicidal or nonsuicidal) |
| SH002 | Personal history of self-harm | Z91.5, Z91.52 (excluding Z91.51, mapped to SA002) | Historical self-harm documentation, intent unspecified |
| SH003 | Self-harm with undetermined intent | Y10-Y19, Y20-Y34; T36-T65 with 6th character "4"; T71 with 6th character "4"; Y87.2 | Episodes where intent is undetermined |

Abbreviations: ICD-10-CM, International Classification of Diseases, Tenth Revision, Clinical Modification; SA, suicide attempt; SH, self-harm; SI, suicidal ideation.
Suicide-related ICD-10-CM codes were reclassified from their original CCSR categories into 6 custom categories to distinguish suicidal ideation, suicide attempts, and self-harm by intent. Poisoning codes were mapped by intent: intentional self-harm to SH001 and undetermined intent to SH003; accidental, assault, adverse effect, and underdosing codes remained in their original CCSR categories. Codes R45.88, Z91.51, and Z91.52, introduced in October 2021, did not appear in the 2016-2020 study dataset but were included in the mapping for future applicability.

**Table S2.** Reclassification of ICD-10-CM Chapter 21/FAC codes.

| **FAC Code** | **CCSR Label** | **Study Decision** | **Note** |
| --- | --- | --- | --- |
| **FAC001** | Encounter for administrative purposes | Exclude | Pure admin contact can lead to detection bias risk; not used as sequence elements. |
| **FAC002** | Encounter for mental health services related to abuse | Include | Clinically relevant psychosocial risk content (abuse-related encounters). |
| **FAC003** | Observation/exam for conditions ruled out | Split (Partial Include) | Split into 4 subgroups: (1) **FAC003_SCREENING** Exclude (utilization); (2) **FAC003_PREGNANCY** Exclude (utilization); (3) **FAC003_ACCIDENT_OBS** Include (21 suicides, post-accident evaluation may capture self-harm); (4) **FAC003_OTHER** → Include (ruled out conditions, observation). |
| **FAC004** | Encounter for prophylactic/other procedures | Exclude | Preventive procedures are utilization, not states. |
| **FAC005** | Encounter for prophylactic measures (excl. immunization) | Exclude | Preventive admin content; not a risk state. |
| **FAC006** | Encounter for antineoplastic therapies | Split (Partial Include) | Split into 2 subgroups: (1) **FAC006_SCREENING** Exclude (cancer screening = utilization); (2) **FAC006_CANCER_HISTORY** Include (Z08, Z483: cancer survivor follow-up encounters, ~23 suicides). |
| **FAC007** | Encounter for mental health conditions | Split (Partial Include) | Split into 2 subgroups: (1) **FAC007_AUTHORITY_EXAM** (Z046) and Include (court-ordered psych exams, 14 suicides, 0.4% rate = HIGH RISK); (2) **FAC007_SCREENING** (Z133X) → Exclude (routine depression/MH screening = utilization). |
| **FAC008** | Neoplasm-related encounters | Split (Partial include) | Split into 2 subgroups: (1) **FAC008_SCREENING** (Z12X) Exclude (not a risk state, cancer diagnosis is captured under NEO category); (2) **FAC008_FOLLOWUP** (Z08, Z483) we include (cancer survivor surveillance). |
| **FAC009** | Implant/device/graft related encounter | Include | Heterogeneous category (orthopedic joints, cardiac devices, catheters, bariatric surgery, etc.); ~650 suicides total; too complex to meaningfully split; keep as composite chronic illness/disability marker. |
| **FAC010** | Other aftercare encounter | Exclude | Broad aftercare = utilization; unless specific suicide-relevant subcode carved out elsewhere. |
| **FAC011** | Counseling related to sexual behavior/orientation | Include | Psychosocially salient for some risk pathways. |
| **FAC012** | Other specified encounters and counseling | Exclude | Generic counseling = utilization/contact. |
| **FAC013** | Contraceptive/procreative management | Exclude | Preventive admin contact; not a risk state. |
| **FAC014** | Medical exam/evaluation | Exclude | Routine checkups inflate contact signal; detection bias. |
| **FAC015** | Resistance to antimicrobial drugs | Exclude | Minimal direct suicide relevance. |
| **FAC0161** | STD - screening & exposure | Include | Psychosocial stressor; stigma/disclosure issues. |
| **FAC0162** | Non-sexual ID - screening & exposure | Exclude | Generic infectious disease screening = utilization. |
| **FAC0163** | Immunization encounter | Exclude | Pure utilization/prevention. |
| **FAC017** | No immunization/under immunization | Exclude | Status markers; not risk states. |
| **FAC018** | Screening for neurocognitive/neurodevelopmental | Exclude | Screening = contact; not a diagnosis state. |
| **FAC019** | Socioeconomic/psychosocial factors | Include | High face-validity psychosocial stressors (housing, employment, relationships, abuse, social isolation); ~100 suicides. |
| **FAC020** | Lifestyle/life management factors | Split | Split into 2 subgroups: (1) **FAC020_TOBACCO** (Z720) Keep (66 suicides, 51% of category, major independent risk factor); (2) **FAC020_OTHER** Keep (high-risk behavior, stress, sleep issues, antisocial behavior; ~64 suicides). |
| **FAC021** | Personal/family history of disease | Include | Personal history of self-harm (Z91.5x) carved into SH/SA002; remaining generic history = moderate yield. |
| **FAC022** | Acquired absence of limb/organ | Include | Chronic disability context (physical loss, body image, functional limitations); ~250 suicides. |
| **FAC023** | Organ transplant status | Include | Major chronic illness context; immunosuppression burden. |
| **FAC024** | Carrier status | Exclude | Low direct suicide relevance. |
| **FAC0250** | Other specified status (catch-all) | Exclude | Generic status codes; moved specific subcategories below. |
| **FAC02501** | Dependency & Assistance Status | Split (6 subgroups) | (1) **FAC02501_DIALYSIS** (67 suicides); (2) **FAC02501_RESPIRATORY** (48 suicides); (3) **FAC02501_MOBILITY** (42 suicides); (4) **FAC02501_OSTOMY** (21 suicides); (5) **FAC02501_ASSISTANCE** (<11 suicides); (6) **FAC02501_OTHER** (46 suicides). All kept as markers of chronic illness burden/dependence. |
| **FAC02502** | Long-Term Medication Use | Split (5) | (1) **FAC02502_ANTITHROMBOTIC** (185 |
|  |  | subgroups + OTHER) | suicides); (2) **FAC02502_DIABETES** (130 suicides); (3) **FAC02502_STEROID** (56 suicides); (4) **FAC02502_PAIN** (30 suicides); (5) **FAC02502_OPIOID** (14 suicides, high clinical relevance); (6) **FAC02502_OTHER** (289 suicides). All kept; reflects chronic disease burden. |
| **FAC02503** | Allergy Status | Split (2 subgroups) | (1) **FAC02503_DRUG** (160 suicides, drug/medication allergies); (2) **FAC02503_NONDRUG** (81 suicides, food/latex/environmental). Dental caries codes (Z9184X) moved to FAC025 catch-all. Both kept. |
| **FAC02504** | Organ Donor Status | Exclude | Administrative marker; not a risk state. |
| **FAC02505** | Patient/Caregiver Non-Compliance | Include | Treatment nonadherence marker (medication, dialysis, dietary); |
| **FAC02506** | Blood Type & Genetic Carrier Status | Split (4 subgroups) | (1) **FAC02506_RH_NEGATIVE** (7 suicides); (2) **FAC02506_RH_POSITIVE** (0 suicides); (3) **FAC02506_GENETIC** (1 suicide, carrier status); (4) **FAC02506_OTHER** (0 suicides). Keep Rh-negative split for biological marker analysis. |
| **FAC02507** | Retained Foreign Body Status | Include | kept for completeness (incidental findings from trauma/procedures). |
| **FAC02508** | Body Mass Index | Split (4 subgroups) | (1) **FAC02508_UNDERWEIGHT** (BMI <20, ~31 suicides, eating disorder marker); (2) **FAC02508_NORMAL** (BMI 20-24.9, ~23 suicides); (3) **FAC02508_OVERWEIGHT** (BMI ≥25, ~74 suicides, depression/metabolic syndrome); (4) **FAC02508_PEDIATRIC** (~3 suicides). All kept; BMI extremes = established suicide risk factors. |
| **FAC02509** | Contact with Hazardous Substances | Include | Occupational/environmental exposure marker. |
| **FAC02510** | Care Decision Status | Split (2 subgroups) | (1) **FAC02510_DNR** (Z66, 49 suicides, 75% of category; end-of-life care marker); (2) **FAC02510_PROCEDURE_NOT_DONE** (21 suicides, treatment refusal/contraindications). Surgical "converted to open" codes (Z533X) moved to FAC0250 catch-all (not care decisions). Both kept. |
| **FAC02511** | Healthcare Access Factors | Include | System barriers, waiting for care, transplant status; 6 suicides but captures access disparities. |
| **FAC02512** | Physical Restraint/Supervision | Include | Acute behavioral crisis markers (Z743: need for supervision, 128 suicides; Z781: physical restraint, 28 suicides); 156 suicides total. |
| **FAC026** | Personal history of nicotine dependence | Include | Z87891 (personal history of nicotine dependence); ~45 suicides; distinct from active F17.X disorder (MBD) and Z720 tobacco use (lifestyle). |
| **FAC027** | Personal history of malignant neoplasm | Include | Z85X codes (personal history of cancer by site); ~256 suicides; distinct from FAC006/FAC008 encounters; kept as composite cancer history marker without subgroups. |
| **FAC028** | Family history of disease | Exclude | Family history codes; genetic risk but not personal illness states. |
| **FAC029** | Genetic susceptibility to disease | Include | Z17X (ER status, 25 suicides) + Z15X (genetic susceptibility, 1 suicide); 26 suicides total; kept as-is (too small to split; ER status dominates). |
| **FAC030** | Personal history of other disease | Include | Heterogeneous catch-all (CVA/TIA, PE, UTI, falls, polyps, TBI, infections, etc.); ~840 suicides across 90+ codes; kept as composite "any significant medical history" marker; too complex to meaningfully split. |

**Table S3.** Discovery cohort characteristics after incidence-density matching.

| Characteristic | Overall (N=7 634) | Suicide (N=694) | Control (N=6 940) | SMD |
| --- | --- | --- | --- | --- |
| Age at index, mean (SD), y | 51.5 (19.4) | 51.5 (19.4) | 51.5 (19.4) | 0 |
| Coverage duration, mean (SD), days | 1 073.9 (445.6) | 1 102.3 (431.1) | 1 071.1 (447.0) | 0.071 |
| Sex, No. (%) |  |  |  |  |
| Male | 5 786 (75.8) | 526 (75.8) | 5 260 (75.8) | 0 |
| Female | 1 848 (24.2) | 168 (24.2) | 1 680 (24.2) | — |
| Encounters, median (IQR) | 37 (15-82) | 32 (12-72) | 38 (15-83) | 0.094 |

Abbreviations: IQR, interquartile range; SD, standard deviation; SMD, standardized mean difference.
Characteristics of the matched cohort used for conditional logistic regression analysis of candidate sequences.
Matching Criteria: Exact match on sex and race/ethnicity; caliper match on age ( $\pm 5$ years); Temporal match requiring controls at-risk and covered at case's index month; Ratio 1:10 (case: control).

**Table S4.** Characteristics of suicide decedents by sequence completion status.

| Characteristic | Any Significant Sequence<br>(N=477) | No Significant Sequence<br>(N=291) | P value |
| --- | --- | --- | --- |
| **Demographics** |  |  |  |
| Age at death, mean (SD), y | 51.9 (19.4) | 49.4 (19.3) | 0.09 |
| Female, n (%) | 137 (28.7%) | 42 (14.4%) | <0.001 |
| White, n (%) | 316 (66.2%) | 146 (50.2%) | <0.001 |
| Black, n (%) | 77 (16.1%) | 42 (14.4%) | 0.59 |
| Asian, n (%) | 20 (4.2%) | 11 (3.8%) | 0.93 |
| Other or unknown, n (%) | 64 (13.4%) | 92 (31.6%) | <0.001 |
| **Insurance Coverage** |  |  |  |
| Medical coverage, n (%) | 477 (100.0%) | 290 (99.7%) | 0.80 |
| Behavioral health coverage, n (%) | 285 (59.7%) | 159 (54.6%) | 0.19 |
| Prescription coverage, n (%) | 449 (94.1%) | 270 (92.8%) | 0.56 |
| **Healthcare Utilization** |  |  |  |
| Total encounters, median (IQR) | 47 (19–94) | 6 (0–22) | <0.001 |
| Unique diagnoses, median (IQR) | 40 (19–82) | 7 (0–19) | <0.001 |
| Inpatient admissions, median (IQR) | 2 (0–7) | 0 (0–0) | <0.001 |
| ED visits, median (IQR) | 1 (0–3) | 0 (0–0) | <0.001 |
| Outpatient visits, median (IQR) | 42 (19–88) | 6 (0–21) | <0.001 |
| Procedures, median (IQR) | 80 (37–171) | 12 (0–44) | <0.001 |

Abbreviations: ED, emergency department; IQR, interquartile range; SD, standard deviation.
Of 768 suicide decedents, 477 (62.1%) completed at least one significant trajectory and 291 (37.9%) did not. Decedents without significant trajectories had substantially lower healthcare utilization across all encounter types, likely reflecting limited clinical engagement or patterns of healthcare-seeking behavior.

**Table S5.** Path-stratified suicide risk after additional adjustment for trajectory duration.

| Trajectory <sup>a</sup> | Path | Events | Model 2 <sup>b</sup> AHR (95% CI) | Model 4 <sup>c</sup> AHR (95% CI) | Model 5 <sup>d</sup> AHR (95% CI) |
| --- | --- | --- | --- | --- | --- |
| Depression → [...] → SI | Other paths | 36 | 1 [Reference] | 1 [Reference] | 1 [Reference] |
|  | Anxiety | 25 | 1.1 (0.6-1.8) | 1.0 (0.6-1.8) | 1.0 (0.6-1.7) |
|  | Anxiety → Anemia | 11 | 4.8 (2.2-10.5) | 4.1 (1.8-9.1) | 6.4 (2.8-14.4) |
| Depression → [...] → Hypertension | Other paths | 51 | 1 [Reference] | 1 [Reference] | 1 [Reference] |
|  | Anxiety | 25 | 1.1 (0.7-1.8) | 1.1 (0.7-1.8) | 1.1 (0.7-1.8) |
|  | Anxiety → Bipolar | 14 | 10.9 (5.7-20.8) | 10.1 (5.2-19.5) | 10.2 (5.3-19.6) |
| Depression → [...] → Hypertension | Other paths | 51 | 1 [Reference] | 1 [Reference] | 1 [Reference] |
|  | Anxiety | 31 | 1.3 (0.8-2.1) | 1.2 (0.8-2.0) | 1.3 (0.8-2.0) |
|  | Anxiety → Hx of self-harm | <11 | 23.3 (10.4-51.9) | 21.6 (9.6-48.4) | 21.9 (9.7-49.3) |
| Depression → [...] → Bipolar | Other paths | 23 | 1 [Reference] | 1 [Reference] | 1 [Reference] |
|  | Anxiety | 22 | 1.4 (0.8-2.6) | 1.4 (0.8-2.6) | 1.5 (0.8-2.7) |
|  | Anxiety → Skin disorders | 11 | 4.3 (2.0-9.4) | 4.6 (2.1-10.2) | 5.2 (2.4-11.4) |
| Depression → [...] → Lipid disorders | Other paths | 45 | 1 [Reference] | 1 [Reference] | 1 [Reference] |
|  | Hypertension | 29 | 0.9 (0.6-1.5) | 0.9 (0.6-1.5) | 0.9 (0.6-1.4) |
|  | Nausea/vomiting | <11 | 2.7 (1.2-5.8) | 2.7 (1.2-5.8) | 2.6 (1.2-5.7) |
|  | Nausea/vomiting → HTN | 12 | 4.7 (2.3-9.3) | 4.7 (2.3-9.4) | 4.4 (2.2-8.9) |
| Anxiety → [...] → SDOH | Other paths | 19 | 1 [Reference] | 1 [Reference] | 1 [Reference] |
|  | SI | 14 | 6.5 (3.2-13.4) | 6.7 (3.2-13.7) | 6.7 (3.2-13.7) |
|  | General symptoms → SI | <11 | 13.4 (5.1-35.6) | 12.3 (4.6-32.8) | 13.6 (5.1-36.5) |
| Depression → [...] → SDOH | Other paths | 16 | 1 [Reference] | 1 [Reference] | 1 [Reference] |
|  | SI | 16 | 5.3 (2.6-11.1) | 5.6 (2.7-11.5) | 5.6 (2.7-11.5) |
|  | General symptoms → SI | <11 | 10.5 (3.9-27.7) | 9.7 (3.6-25.6) | 11.4 (4.3-30.5) |

Abbreviations: AHR, adjusted hazard ratio; CI, confidence interval; HTN, hypertension; Hx, history; SI, suicidal ideation; SDOH, social determinants of health.
<sup>a</sup> Trajectories match those in Table 3. Within each block, patients who completed the same start → end transition were stratified into mutually exclusive paths. The reference group includes patients who reached the endpoint through paths not explicitly modeled.
<sup>b</sup> Model 2 (primary): age, sex, race/ethnicity, strata (entry year), observable duration, 12-month coverage completeness, Charlson Comorbidity Index at trajectory start. Reproduced from Table 3 for comparison. <sup>c</sup> Model 4: Model 2 + trajectory duration in years. Trajectory duration was defined as the time from first to last diagnosis in the sequence and was added as a covariate to assess whether the speed of diagnostic accumulation confounded path-specific associations.
<sup>d</sup> Model 5: Model 3 (sensitivity: Model 2 with Charlson Comorbidity Index at trajectory end) + trajectory duration in years.

